# Predicting Atrial Fibrillation Ablation Outcomes: A Machine Learning Approach Leveraging a Large Administrative Claims Database

**DOI:** 10.1101/2024.11.16.24317420

**Authors:** Yijun Liu, Mustapha Oloko-Oba, Kathryn Wood, Michael S. Lloyd, Joyce C. Ho, Vicki Stover Hertzberg

## Abstract

**Background:** Atrial fibrillation (AF) ablation is an effective treatment for reducing episodes and improving quality of life in patients with AF. However, in some patients there are only modest long-term AF-free rates after AF ablation. There is a need to address the limited benefits some patients experience by developing predictive algorithms to improve AF ablation outcomes.

**Objective:** The authors aim to utilize machine learning models on claims data to explore if innovative coding models may lead to better patient outcomes than use of traditional stroke risk score prediction.

**Methods:** The Merative MarketScan® Research Medicare data was used to examine claims for AF ablation. To predict 1-year AF-free outcomes after AF ablation, logistic regression and XGBoost models were used. Model predictions were compared with established risk scores CHADS_2_ and CHA_2_DS_2_-VASC. These models were also assessed on subgroups of patients with paroxysmal AF, persistent AF, and both AF and atrial flutter from October 2015 onwards.

**Results:** The sample included 14,521 patients with claims for AF ablation. XGBoost achieved an area under the receiver operating characteristic curve (AUC) of 0.525, 0.521, and 0.527 for the entire AF ablation population, female, and male, respectively. Within the subgroups, machine learning models performed the best for the paroxysmal AF subgroup using ICD codes, demographic information, and comorbidity indexes, achieving an AUC of 0.546.

**Conclusion:** Machine learning models outperformed CHADS_2_ and CHA_2_DS_2_-VASC in all AF ablation patient groups (whole population, female, and male). Using patient data for those who had their AF ablation on or after October 2015, machine learning models performed best in all subgroups and the population, indicating that including ICD codes in machine learning models may improve performance.

## Introduction

Affecting over 6 million people in the U.S., atrial fibrillation (AF), the most common cardiac arrhythmia, is a major public health concern that will continue to increase with the aging United States (US) population.^1–4^ AF is costly to health care systems and leads to significant health consequences of stroke, heart failure, dementia, and decreased quality of life.^5^ Although there is no cure for AF currently, AF ablation is the most effective treatment to restore normal sinus rhythm and decrease symptoms in episodes of paroxysmal or persistent AF, thereby reducing AF burden and improving quality of life.^1–3^ AF is associated with increased risk of cardiovascular events that may affect treatment outcomes, yet there limited knowledge of personalized risk stratification for patients undergoing AF ablation. Existing risk scores, like CHADS_2_ and CHA_2_DS_2_-VASc, have traditionally been applied to predict stroke risk and are now also utilized in risk assessment following COVID-19, heart surgery, and AF ablation.^6–13^

Success rates for AF ablation from the literature vary based on individual clinical variables such as type of AF, left atrial size or volume index,^1,2,4^ yet these variables are frequently difficult to access in large electronic health record datasets (EHR). Patients continue to experience episodes of AF following an initial AF ablation with long-term AF-free rates after de novo AF ablation reported as 50% to 75%.^1,2,14^ Additionally, the chances of developing any complications after AF ablation range around 6%, with 0.1% to 0.9% of patients experiencing complications that could result in death.^15–17^ Given the modest success rates of AF ablation, prediction of outcomes could be personalized to more easily identify those who would be most likely to benefit from an AF ablation.

Machine learning (ML) has emerged as a powerful approach that leverages increased computational power with large datasets to help achieve complex decisions to guide clinical practice.^18^ Artificial intelligence and ML have been used in the field of electrophysiology since the 1970s for automated ECG interpretation.^18,19^ More recently, innovations in algorithms, development and labeling of large databases, and improvements in hardware and software have rapidly increased the role of ML in cardiac electrophysiology and cardiovascular imaging to identify predictors of patient outcomes.^20^ ML has already been used to improve prediction of AF ablation outcomes, primarily via electronic health records (EHRs). Nevertheless, health systems are not widely interoperable,^21^ making it both costly and challenging to extend these prognostic tools across multiple health systems. Studies utilizing EHR data have often been limited to datasets from one to two hospitals, limiting the generalizability of the models and hindering broad adoption.^22,23^ Claims data, on the other hand, is commonly collected, more readily available, and usually collected on a large national scale.^21^ Although EHR data, which can include medications, laboratory data, and radiology reports, is more granular than claims data and can offer more accurate predictions, claims-based prognostic models can offer better scalability across health systems and populations.^24^

In this study, we propose to develop ML-based predictive models for outcomes of de novo AF ablation procedures using national level claims data in the United States. Our goal is to evaluate a ML derived risk prediction model for AF ablation patient outcomes. We hypothesize that the ML models will be comparable to or exceed existing AF risk scores with respect to predictive power. Existing risk scores including CHADS_2_ and CHA_2_DS_2_-VASc have achieved non-trivial improvements in predicting the outcomes of AF procedures, achieving area under the receiver operating characteristic curve (AUROC) of 0.785 and 0.830 respectively in a dataset consists of 565 patients.^25^ Thus, in this study, we utilize CHADS_2_ and CHA_2_DS_2_-VASc as a baseline to compare with our ML approaches. We will also characterize outcomes by subgroups as well as examine differences in AF ablation outcomes by sex.

## Methods

This research leveraged de-identified claims data sourced from the Merative MarketScan® Research Medicare Databases (MarketScan, Inc., Ann Arbor, MI) to investigate AF ablation (CPT code ‘93656’) claims spanning January 1, 2013, to December 31, 2020. MarketScan contains claims made for individuals with Medicare Supplemental and Medicare Advantage plans.

Inclusion criteria comprised patients across inpatient admission (I), inpatient services (S), and outpatient services (O) tables in the database. Medical history and postoperative outcomes were scrutinized using claims data from January 1, 2011, to December 31, 2021, with a focus on patients possessing valid patient IDs for traceability. Each patient’s medical history included all ICD codes from visits within the two years before the initial occurrence of AF ablation. The final cohort comprised 14,521 patients after excluding those without valid patient IDs. The research was conducted at Emory University, Atlanta, GA.

### Patient Population

We included patients undergoing AF ablation (CPT code ‘93656’) between January 1, 2013, and December 31, 2020, using the I, S, and O tables from Medicare claims. Only the I, S, and O tables have ICD codes which are the primary factors we used for our predictions. These patients were validated to avoid CPT coding errors by verifying the co-presence of AF (ICD-9 code of ‘427.31’ or ICD-10 code of ‘I48.X’) and AF ablation.

The decision to include exclusively Medicare patients stemmed from the MarketScan database limitations preventing integration of commercial and Medicare claims, as the unique patient IDs differ between the Medicare and commercial claims datasets. Additionally, mean age for first AF ablation usually happens around 55-62.^2–4^ Medicare therefore covered a greater range of aged patients in the database. Moreover, the absence of postoperative outcomes for numerous patients in the commercial database rendered it unsuitable for this study.

### Definition

Our study’s objective is to forecast the binary outcome—success or failure—of AF ablation prior to the procedure based on patient past medical history and demographics. Success entailed the absence of recurring AF or AF ablation within the 6-month interval from the end of the sixth to the end of the twelfth month following the initial AF ablation procedure. Conversely, failure was defined by any occurrence of recurring AF or AF ablation during the same 6-month period. The patient population was further validated by confirming that they had revisited the clinics within a year after ablation to ensure the success population was a true success.

While the O datasets comprehensively documented the operation date for AF ablation, the I and S datasets exclusively provided admission and discharge dates. In navigating this constraint, we pragmatically designated the admission date from the I and S datasets as a surrogate for the AF ablation operation date in our analysis. This procedural adjustment was necessitated by the dataset structure, allowing us to maintain temporal coherence in our predictive modeling.

The study employed subgroup analysis, where we identified groups with paroxysmal AF, persistent AF, and AF patients with flutter. ICD-10 codes clearly distinguish between paroxysmal AF, persistent AF, and chronic AF, in comparison to ICD-9 codes that do not make these distinctions. Within the Medicare datasets, all patient records after October 1, 2015, adopted ICD-10 codes instead of ICD-9 codes. To reflect current changes in terminology of types of AF, we combined persistent AF and chronic AF in the database as persistent AF. We defined paroxysmal AF as patients with ICD-10 code of I48.0, I48.20, and free of persistent AF code; persistent AF as patients with I48.1, I48.11, I48.19, I48.2, and I48.21. Patients with atrial flutter had any AF code with I48.3 (ICD-10) or I48.4 (ICD-10) or 427.32 (ICD-9).

### Data Processing

Each patient was distinctly identified by a unique patient ID that facilitates cross-referencing across datasets, namely the I, S, and O tables, housing medical claims for individual medical visits. Our predictive focus necessitated a retrospective analysis, limited to the patient’s history preceding the date of AF ablation.

To present a comprehensive historical snapshot, we constructed a table encompassing patient ID, demographic details (sex, region, age, and industry), the earliest AF ablation date, the failure date (if applicable), and an exhaustive list of ICD and CPT codes from past medical visits. This retrospective tableau extended back over a 2-year period.

In adapting to Medicare’s transition from ICD-9 to ICD-10 post-October 2015, we integrated the conversion from ICD-10 to ICD-9 codes using the ICD-10 Lookup tool.^26^ To simplify our dataset and maintain a manageable feature set, we retained only the first three digits of the converted ICD codes.

Within the claims data, we calculated two established indices—the Charlson Comorbidity Index (CCI) and the Elixhauser Comorbidity Index (ECI)—to capture patients’ comorbid conditions.^27,28^ These indices used a weighted system based on specific conditions to provide a score, with higher values indicating more severe comorbidities.

### Modeling

We used two popular and efficient supervised ML classifiers: Logistic Regression and XGBoost (eXtreme Gradient Boosting) (XGB).^29^ Logistic Regression computes the probability of a binary outcome by employing a logistic function (sigmoid curve) to transform the linear combination of input features into probabilities. This model is particularly advantageous due to its simplicity and interpretability, especially in scenarios where the relationship between input variables and the outcome is expected to be linear. On the other hand, XGB represents a more sophisticated approach. As an ensemble technique, it constructs multiple decision trees in a sequential manner, with each subsequent tree focusing on addressing the errors made by its predecessors. This method does not presuppose a linear relationship between input and output variables, offering greater flexibility and efficacy in dealing with larger and more intricate datasets. Despite its computational intensity, XGB is celebrated for its high efficiency and versatility, making it a potent tool in predictive modeling, especially in situations where the complexity of the data surpasses the capabilities of simpler models like Logistic Regression.^24^

The CHADS_2_ and CHA_2_DS_2_-VASC risk scores have been widely used to predict stroke risk in patients with AF.^12,13,25^ These risk scores more recently have been used to predict outcomes in patients with AF, heart failure, coronary artery disease, and postoperative AF undergoing cardiovascular surgical procedures.^6,11^ Still other investigators have reported that the CHADS_2_ and CHA_2_DS_2_-VASC risk scores were useful predictors of adverse events after AF ablation.^7^ Therefore, we chose to use CHADS_2_ and CHA_2_DS_2_-VASC risk score as our baseline comparison.

### Statistical Analysis

For the continuous variable age, we used a t-test to obtain the p-value. For categorical variables, we used Chi-square tests. Continuous variables were reported as the mean ± standard deviation, while categorical variables were expressed as percentages. The area under receiver operating characteristic curve (AUC) was used to assess the performance of each model.

For two-point statistics, bootstrap resampling was used to generate a distribution and then perform a t-test to obtain the p-values. Specifically, bootstrap resampling was only used to obtain the p-values in table 2.

### Ethical Considerations

This study used commercially available data that have been deidentified. As such, the study was deemed exempt by Emory University Institutional Review Board.

## Results

The demographic and clinical profiles of patients with AF are detailed in Tables 1 and 2. Our study cohort consisted of 14,521 patients, with an average age of 71.5 years (SD = 5.31). A successful outcome from AF ablation procedures was observed in 54.01% (n=7,843) of patients. Females constituted 39.94% of the study population. Clinically, 24.73% (n=3,591) of the patients were diagnosed with concomitant atrial flutter. The precise identification of patients with paroxysmal and persistent AF was limited, relative to the total cohort, due to the use of ICD-9 instead of ICD-10 prior to October 2015. A total of 7,646 patients were identified using ICD-10 codes for AF ablation, demonstrating a slightly reduced AF ablation success rate of 53.28% in comparison to the broader patient population. A subset of 6,983 patients was accurately categorized as having paroxysmal or persistent AF. Within this subset, 37.63% (n=2,877) were diagnosed with paroxysmal AF, while 53.70% (n=4,106) had persistent AF. The AF ablation success rates for paroxysmal and persistent AF were 52.55% and 53.90%, respectively.

**Table 1.**
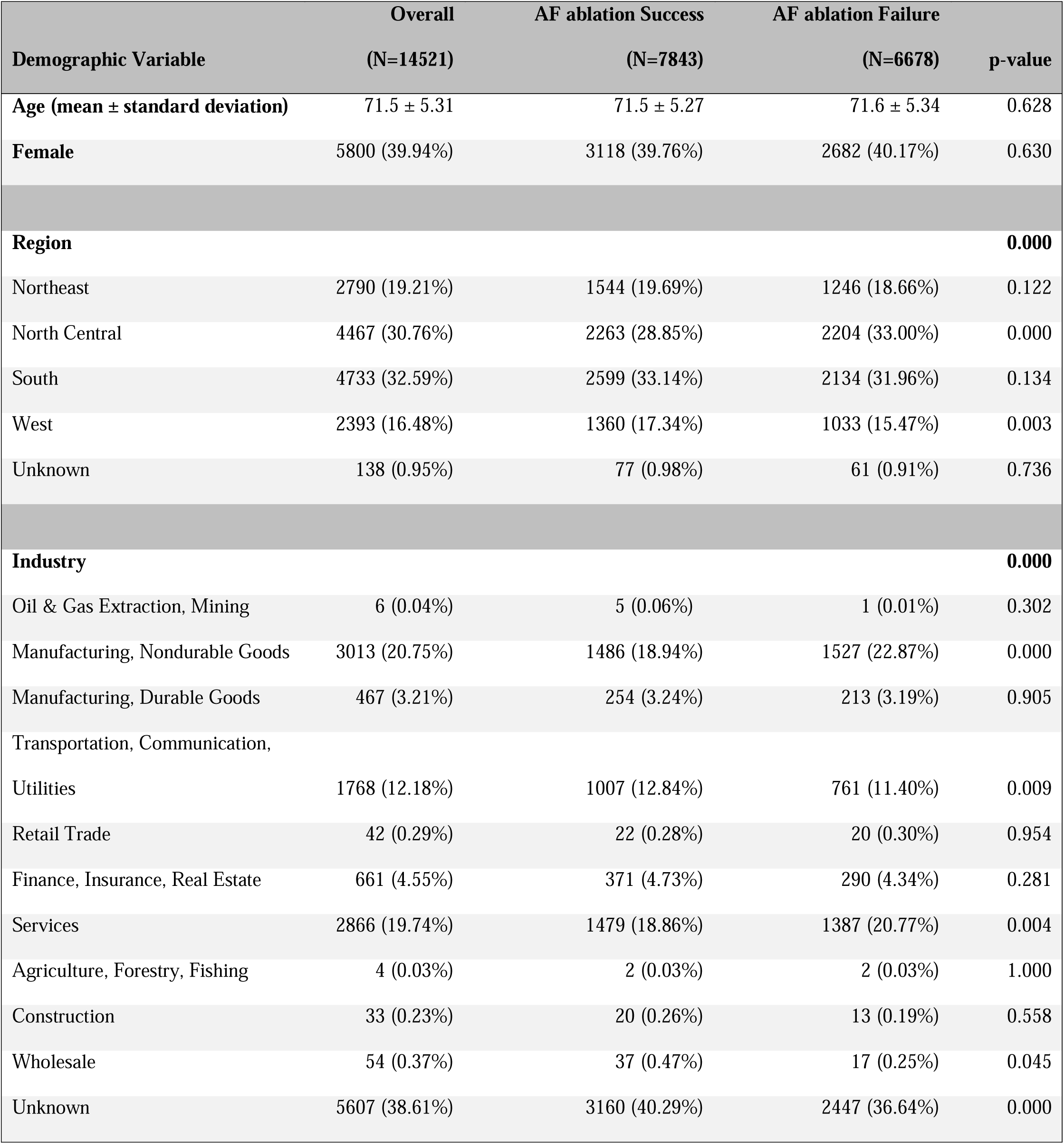
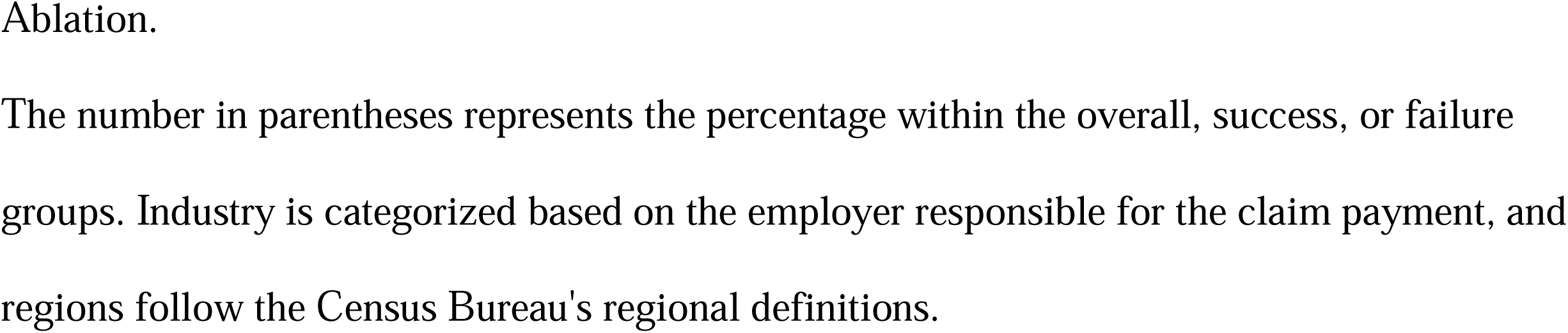
Baseline Demographic Characteristics of Patients undergoing Atrial Fibrillation.

**Table 2.**
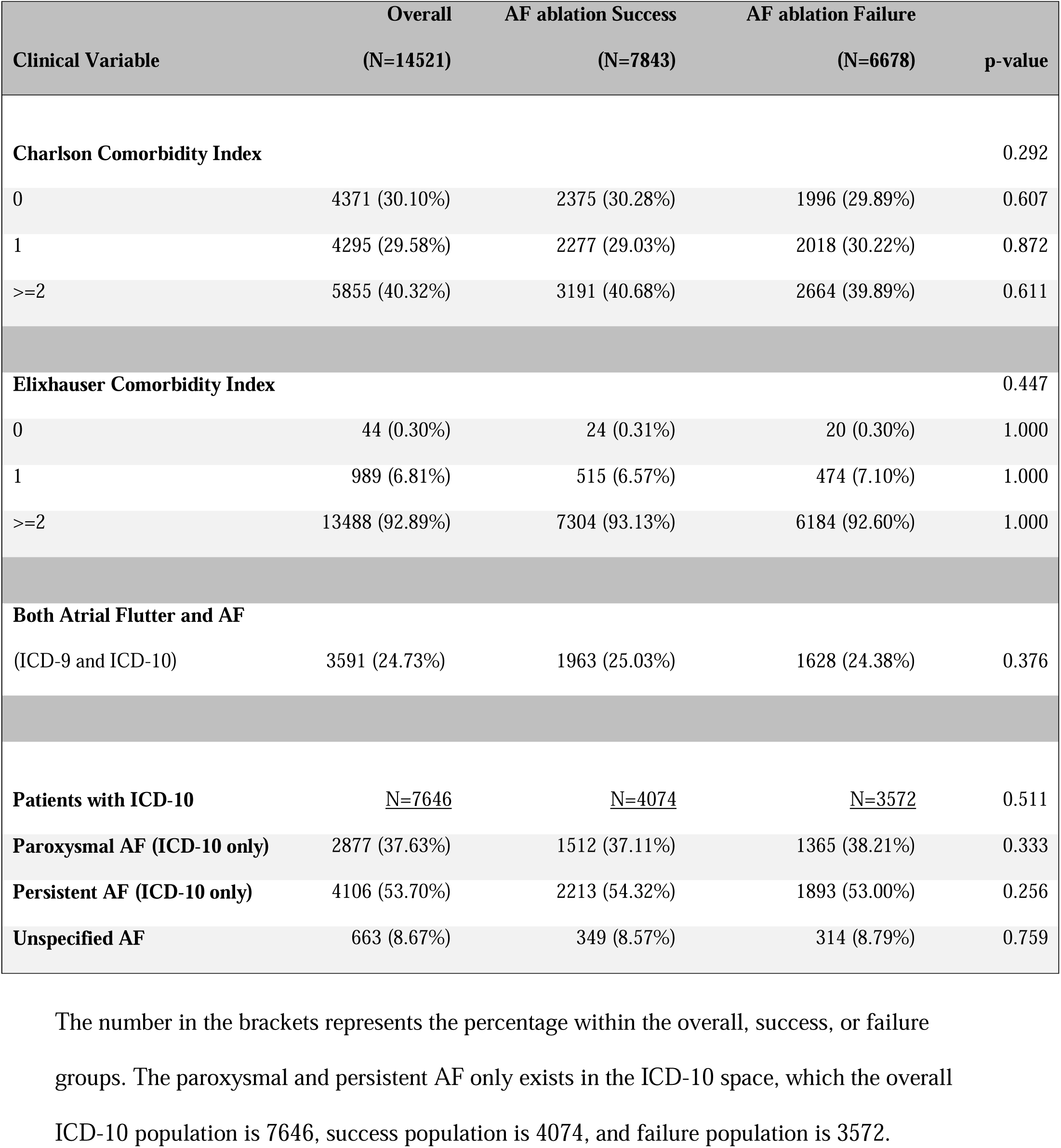
Baseline clinical characteristics of patients in sample undergoing AF ablation.

Table 3 showed the comparative performance, as measured by the Area Under the Receiver Operating Characteristic Curve (AUC ROC), of the XGB machine learning model and the risk scores, which were CHADS_2_ and CHA_2_DS_2_VASc, in predicting the outcomes of AF ablation. CHADS_2_ and CHA_2_DS_2_VASc are stroke prevention risk scores, with CHADS_2_ involving congestive heart failure, hypertension, age ≥75, diabetes, stroke (doubled) and CHA_2_DS_2_VASc involving congestive heart failure, hypertension, age ≥75 (doubled), diabetes, stroke (doubled), vascular disease, age 65 to 74 and sex category (female). Notably, both CHADS_2_ and CHA_2_DS_2_VASc scores yielded predictions worse than random chance (AUC ROC < 0.5) except for CHA_2_DS_2_VASc in female group (which ROC = 0.500, equaled to the chance of random guessing). The XGB model exhibited modest predictive accuracy across the entire study population with an AUC ROC of 0.525. This accuracy was slightly improved (0.527) for males but marginally decreased (0.521) for females. Nevertheless, the XGB model surpassed the predictive capabilities of the CHADS_2_ and CHA_2_DS_2_VASc scores across all patient cohorts.

**Table 3.** Performance Comparison between XGB and CHADS_2_ and CHA_2_DS_2_VASc Risk Scores Stratified by Sex).

|  | **XGB**<br>**(mean ± SD)** | **CHADS<sub>2</sub>**<br>**Scores** | **CHA<sub>2</sub>DS<sub>2</sub>VASc**<br>**Scores** | **p-value**<br>**(XGB C2)** | **p-value**<br>**(XGB C2V)** | **p-value**<br>**(C2 C2V)** |
| --- | --- | --- | --- | --- | --- | --- |
| **Population**<br>**(n=14,521)** | 0.525<br>(0.522-0.527) | 0.498 | 0.498 | **0.000** | **0.000** | 0.507 |
| **Female**<br>**(n=5,800)** | 0.521<br>(0.517-0.525) | 0.498 | 0.50 | **0.000** | **0.000** | 0.510 |
| **Male**<br>**(n=8,721)** | 0.527<br>(0.524-0.531) | 0.498 | 0.498 | **0.000** | **0.000** | 0.498 |

All performance values are reported in terms of AUC ROC. Cell values for XGB report average ± standard deviation (SD).

Table 4 presented type of atrial arrhythmia subgroup analyses focusing on patients with ICD-10 codes (patients’ AF ablation dates were on or after October 2015). ICD-10 codes are more specific and enables patients to be identified as having paroxysmal AF or persistent AF. In table 4, we presented results for patients with paroxysmal AF, persistent AF, and those who had both AF and atrial flutter (in ICD-10 space), utilizing three distinct datasets: 1) comprising converted, simplified 3-digit ICD codes, demographic information, and comorbidity scales (Charlson Comorbidity Index and Elixhauser Comorbidity Scale); 2) demographic data and comorbidity scales; and 3) solely demographic information. Across all subgroups and the entire ICD-10 population, models based on the first dataset type demonstrated superior predictive performance. The highest AUC ROC was observed in the entire ICD-10 population, achieving 0.550. Among subgroups, the highest AUC ROC was found in patients with paroxysmal AF (0.546) when ICD codes were included. For the first dataset type, the AUC ROC values for paroxysmal AF, persistent AF, AF patients with atrial flutter, and the entire population were 0.546, 0.526, 0.521, and 0.550, respectively. The lowest AUC ROC in each subgroup occurred for AF patients with atrial flutter using only demographic data (0.511). Similarly, for paroxysmal AF and the entire ICD-10 population, the lowest AUC ROC (both 0.534) was observed using demographic data alone. In contrast, the lowest AUC-ROC for persistent AF (0.516) was observed when using demographics combined with comorbidity scales.

**Table 4.** ROC Performance by Clinical and Demographic Predictors Across Atrial Arrhythmia Subgroups.

|  | **ICD+Demo+**<br>**(CCI+ECI)** | **Demo+ (CCI+ECI)** | **Demo** |
| --- | --- | --- | --- |
| **Paroxysmal AF** | 0.546 | 0.543 | 0.534 |
| **(n=2,877)** | (0.542-0.551) | (0.538-0.549) | (0.527-0.540) |
| **Persistent AF** | 0.526 | 0.516 | 0.525 |
| **(n=4,106)** | (0.520-0.532) | (0.512-0.520) | (0.520-0.530) |
| **ICD-10, With Atrial**<br>**Flutter** | 0.521 | 0.514 | 0.511 |
| **(n=1,503)** | (0.514-0.530) | (0.504-0.525) | (0.504-0.519) |
| **ICD-10 Population** | 0.550 | 0.536 | 0.534 |
| **(n=7,646)** | (0.545-0.554) | (0.533-0.541) | (0.531-0.537) |

This population only includes patients who had their first AF ablation in or after October 2015. “ICD” means ICD codes of patients past medical history; “Demo” represents demographic variables (region, sex, age, and industry); the numbers in the brackets represent the confidence interval after 30 Monte Carlo runs.

## Discussion

In this study, we have developed ML learning models that predict outcomes of de novo AF ablation procedures. Our XGB model demonstrated improved performance and predictive power as compared to CHADS_2_ and CHA_2_DS_2_VASc in all patient and sex subgroups. However, XGB’s predictive ability of outcomes after AF ablation was found to be lower in females than it was in males or in the entire population. There was no difference in predictive power when comparing CHADS_2_ to CHA2DS2-VASc risk scores for outcomes after AF ablation except for females where CHA_2_DS_2_-VASc performs better than CHADS_2_. Additionally, in comparing outcomes in patient subgroups of types of AF (paroxysmal, persistent, or AF with atrial flutter) the ICD codes model demonstrated better prediction power than using either only demographic/clinical variables or those variables plus comorbidity scales, the Charlson Comorbidity Index (CCI) and the Elixhauser Comorbidity Index (ECI). Use of these machine learning models may be useful in clinical practice in patient selection for AF ablation in the future.

Previous clinical models for predicting AF ablation success have reported an AUC ranging from 0.55 to 0.65, with only three models achieving an AUC of 0.75.^8–10^ In other studies, CHADS_2_ and CHA_2_DS_2_-VASc can usually achieve an AUC of 0.785 and 0.830 respectively in predicting AF ablation.^25^ However, in our study, CHADS_2_ and CHA_2_DS_2_-VASc only achieved an AUC of 0.498-0.5, performing mostly worse than random guessing (AUC = 0.5). This finding highlights the difficulty in predicting the success and failure of AF ablation for the dataset used in this study. In contrast, our machine learning (ML) models achieved an AUC of 0.521-0.527, outperforming both CHADS_2_ and CHA_2_DS_2_-VASc with statistical significance (p-value = 0.000) in all patient groups (whole population, female, and male) for this dataset.

In addition to demonstrating that ML models outperform these risk score predictions, we have also conducted an analysis to understand what types of features should be included in the ML models. We identified three sets of features and three clinically meaningful subgroups in the population (and the whole population). The three sets of features respectively were: 1) comprised with ICD codes, demographic information, and comorbidity scales; 2) comprised with demographic data and comorbidity scales; and 3) restricted to demographic information; and the three clinically meaningful subgroups are: paroxysmal AF, persistent AF, as well as AF patients with atrial flutter. These subgroups could only be identified in the ICD-10 space, and thereby we also compared these subgroup results with the ICD-10 population. This resulted in 16 unique ML models for each combination of feature sets and subgroups. Across all subgroups, ML models performed best when including ICD codes as features, indicating that the incorporation of ICD codes improved model performance. Among the three subgroups (paroxysmal AF, persistent AF, and patients with atrial flutter), the ML models performed best for paroxysmal AF patients and AF patients with atrial flutter have the least success. The entire ICD-10 population achieved the highest overall AUC ROC compared to other subgroups, which was likely due to the larger sample size.

Our findings demonstrate that ML models using ICD codes to estimate AF ablation procedural outcomes are robust and valid across all populations. Improvement of outcome predictions for AF ablation using ML has the potential for widespread use in research and clinical practice to determine optimal patient selection for AF ablation and AF patient management. Advances in artificial intelligence and ML technology have an ability to rapidly analyze and synthesize innumerable variables to predict outcomes of AF ablation and discover new patterns of clinical variables that greatly surpass prior conventional methods of gauging success. These findings will be important to consider as healthcare policymakers struggle to allocate limited resources to as many patients as possible and search for ways to improve patient outcomes. ML technologies will play increasingly more important roles in medicine with future advances and as we better learn how to incorporate ML for improvements in clinical practice and patient outcomes.

### Limitations

This data used in this study comprised health insurance claims data filed for people holding Medicare Advantage or Medicare Supplemental plans. As with all administrative data, some human errors may be present. We addressed these issues by reporting clear descriptions of our data source, the inclusion and exclusion criteria, the billing codes used, and any potential confounding factors included in the modeling. We also verified the patients with AF ablation by identifying at least one past occurrence of AF in their past medical history.

## Data Availability

The machine learning models produced in the present study are available upon reasonable request to the authors. The code is also publicly released in a GitHub repository.

https://github.com/isSherrrrry/AFA-Claims-CodeRelease

## Abbreviations

AF: Atrial fibrillation
AUC: Area under the curve
CCI: Charlson comorbidity index
Com: Comorbidity Indices
Demo: Demographic characteristic(s)
ECG: Electrocardiogram
ECI: Elixhauser comorbidity index
EHR: Electronic health record(s)
I Table: Inpatient Admission Table
ICD: International Classification of Disease
ML: Machine learning
O Table: Outpatient Services Table
ROC: Receiver operating characteristic
S Table: Inpatient Services Table
SD: Standard Deviation
US: United States
XGB: XGBoost
XGBoost: Extreme Gradient Boosting

## Acknowledgment

This research is supported by R21HL156184.

No generative AI models (including Large Language Models such as ChatGPT) were used in the conduct of this research nor in the writing of this manuscript. We use python scikit-learn module to perform logistic regression and XGB which formed the body of this research.^30^

## Supplemental Materials

The code used for this study is publicly released in the GitHub Repository: https://github.com/isSherrrrry/AFA-Claims-CodeRelease

## Notes

**Funding:** This work was supported by grant R21HL156184 (PI: Vicki Stover Hertzberg) from the National Institutes of Health to Emory University, Atlanta, Georgia.

### Competing Interest Statement

The authors have declared no competing interest.

### Funding Statement

This study was supported by the National Institute of Health / National Heart, Lung, and Blood Insitute award R21HL156184.

### Author Declarations

Emory University's Institutional Review Board waived ethical approval for this work.

### Summary of Updates

Updated details for the ICD-10 modeling assessments.

